# Early Detection of Lung Cancer in the NLST Dataset^*^

**DOI:** 10.1101/2023.03.01.23286632

**Authors:** Pritam Mukherjee, Anna Brezhneva, Sandy Napel, Olivier Gevaert

## Abstract

Lung Cancer is the leading cause of cancer mortality in the U.S. The effectiveness of standard treatments, including surgery, chemotherapy or radiotherapy, depends on several factors like type and stage of cancer, with the survival rate being much worse for later cancer stages. The National Lung Screening Trial (NLST) established that patients screened using low-dose Computed Tomography (CT) had a 15 to 20 percent lower risk of dying from lung cancer than patients screened using chest X-rays. While CT excelled at detecting small early stage malignant nodules, a large proportion of patients (*>* 25%) screened positive and only a small fraction (*<* 10%) of these positive screens actually had or developed cancer in the subsequent years. We developed a model to distinguish between high and low risk patients among the positive screens, predicting the likelihood of having or developing lung cancer at the current time point or in subsequent years non-invasively, based on current and previous CT imaging data. However, most of the nodules in NLST are very small, and nodule segmentations or even precise locations are unavailable. Our model comprises two stages: the first stage is a neural network model trained on the Lung Image Database Consortium (LIDC-IDRI) cohort which detects nodules and assigns them malignancy scores. The second part of our model is a boosted tree which outputs a cancer probability for a patient based on the nodule information (location and malignancy score) predicted by the first stage. Our model, built on a subset of the NLST cohort (*n* = 1138) shows excellent performance, achieving an area under the receiver operating characteristics curve (ROC AUC) of 0.85 when predicting based on CT images from all three time points available in the NLST dataset.

## 1. Introduction

The National Lung Screening Trial (NLST) [9] screened more than 50,000 high risk patients between 55 and 74 years of age with long smoking history and showed that patients screened with low-dose CTs had 15 to 20% lower risk of dying from lung cancer than patients who were screened using chest X-rays [8, ^*\**^ Supported by NIH grants U01 CA1964050 and U01 CA187947. 10]. Although this resulted in a higher false positive rate (*>* 90%) in the NLST, complications from invasive diagnostic evaluation procedures were uncommon, leading to better overall outcomes for participants.

The high false positive rate (with corresponding low positive predictive values) does, however, lead to more diagnostic procedures, biopsies and other invasive procedures. This raises the cost as well as the risk of over-diagnosis. The recent advent and successes of quantitative imaging [5, 6] presents a tempting opportunity: quantitative imaging or radiomics may be able to identify and leverage subtle morphological features of the lung nodules and/or the surrounding tissue to assess the risk of lung cancer, and provide a cost-effective and non-invasive alternative to other commonly used followup procedures.

Using a conventional radiomics approach to tackle this issue, however, poses significant challenges, including the unavailability of nodule segmentation for the NLST cohort, which is required for a radiomics approach to be successful. Convolutional Neural Networks (CNN) on the other hand have proven very effective in various image-based tasks including lung nodule segmentation [12] and lesion detection[13, 7]. They do not require handcrafted features and, given enough data, can automatically extract high-level abstract features and capture non-linear classification boundaries.

In this work, we used a two-stage model to predict if a patient with lung nodules is likely to develop cancer in the future based on their lung CTs only. In the context of the NLST, we considered patients who screened positive due to the presence of lung nodules *>* 4 mm in diameter, and trained a model to predict whether they had or would develop cancer in the subsequent years. We used a CNN in the first step in order to detect nodules and predict malignancy, and used a boosted tree in the second stage to predict per-patient risk of cancer based on the nodule information in the first stage. The second stage also takes into account available longitudinal data from previous time-points while making predictions. Our model shows promising predictive performance when predicting on CT images from all three time-points available in the NLST dataset.

## 2 Materials and Methods

### 2.1 Overview of the NLST dataset

The NLST [9] is a randomized, multi-site trial conducted in collaboration between Lung Screening Study (LSS) and American College of Radiology Imaging Network (ACRIN), that examined lung cancer-specific mortality among participants in a high-risk cohort. The aim was to determine whether screening for lung cancer with low-dose helical computed tomography (CT) reduces mortality from lung cancer in high-risk individuals relative to screening with chest radiography. To that end, over 53,000 high risk individuals between 55 and 74 years of age with heavy smoking history were enrolled between 2002 and 2004, and approximately half of them were screened with low-dose CT, while the remaining were screened with chest X-rays. Each participant underwent three annual screenings from 2002–2007, with follow-up post-screening through 2009.

Out of 26732 patients in the low-dose CT arm of the study, cancer was confirmed in only 1083 (*≈* 4%) patients during the course of the study. On the other hand, at each of three screening time-points, say T0, T1, and T2, respectively, a relatively large proportion–27%, 26% and 16%–of patients screened positive, based on the presence of non-calcified nodules or masses *≥*4 mm in diameter or any other abnormalities judged suspicious for lung cancer by the radiologist. About 9%, 6% and 8% of the patients who screened positive at time-points T0, T1, and T2, respectively, were confirmed with cancer either during screening or the subsequent follow-up within the duration of the study.

### 2.2 Study Cohort

For this study, the patients were chosen from the NLST as follows: In the CT arm of NLST (*n* = 26732), 7191 patients screened positive at T0. Most of them (*n* = 7096) had lung nodules greater than 4 mm in diameter. Let us call this set of patients *P* Out of *P*, only 620 patients were diagnosed with cancer at either T0, or T1, or T2 or in the subsequent years while which the patients were followed. We considered these 620 cancer patients as the *cancer-positive* cohort. Next, we selected 620 patients from *P* who were never diagnosed with cancer during the duration of the study and were demographically matched with the cancer-positive cohort, defined as the *control* cohort. The demographic matching was implemented as follows: for each patient in the case cohort, we picked one patient from *P* with the matching gender, and at minimum distance (in the *l*_2_ sense) with respect to age, smoke-years and pack-years (the number of packs per day times the number of years smoked). The resulting control cohort was well-matched with respect to race and ethnicity as well. Finally, we excluded patients for whom either CT scans were unavailable for any time point or if the slice thickness was larger than 3 mm for any of the time points, and ended up with a total of 1138 patients comprising 553 cancer patients and 585 no-cancer.

### 2.3 Available Data

For the selected case and control cohorts, CT imaging data for all three screening time-points was downloaded from The Cancer Imaging Archive (TCIA)[3] with approval from the National Cancer Institute. We also downloaded additional clinical data including demographic data, as well as lung cancer data such as the time of cancer confirmation and size of the lesion at the time of diagnosis. Precise location information for the detected lung nodules was, however, unavailable.

### 2.4 Learning Model

#### Model Description

Our model has two stages: 1. a CNN model for generating lung nodule candidates and assigning them malignancy scores, and 2. a gradient boosted tree that combines the nodule information output by the first stage, for each patient and outputs a probability of cancer.

The architecture of the CNN model in the first stage is similar to the architecture adopted in [11, 4]. Essentially, it has five sequential convolutional layers (Conv3D), each with ReLU activation and followed by a 3D MaxPool layer. The output of the fifth convolution block is fed to two branches: one for predicting nodule probability and the other for predicting nodule malignancy. The branch predicting nodule probability is a convolutional layer with sigmoid activation function, while the branch for malignancy prediction is a convolutional layer with ReLU activation, representing the predicted malignancy score. The input to model is a 3D patch 32 × 32 × 32 pixels in size. Nodule probability represents the probability that the input 3D patch contains a nodule. The second output, malignancy is the predicted ‘malignancy score’ of the nodule. Given a preprocessed (Section 2.4) CT volume, 3D patches of 32 × 32 × 32 pixels are extracted using a sliding window and fed to the CNN model. If the predicted nodule probability is greater than a threshold (chosen to be 0.6 in our case), the location of the patch, along with the predicted nodule probability and predicted malignancy is recorded and used for cancer risk prediction in the second stage. The second stage of our model consists of a xgboost[2] classifier. For each CT scan, the input feature set comprises the number of predicted nodule candidates (nodule probability *>* 0.6) as well as the candidate location (x, y and z coordinates), nodule probability and predicted malignancy for the top *k* = 5 candidates with highest predicted malignancy. We built three different stage 2 classifiers: the first for predicting cancer using T0 scans, the second using T0 and T1 scans and the third using T0, T1 and T2 scans. When using scans from more than one time-point, we picked one CT volume uniformly at random (if multiple volumes were present) from each time-point and concatenated the corresponding input feature sets. The classifier was trained using binary labels: cancer vs no-cancer. The output represents the probability of cancer for the patient.

#### Training Data

The CNN model in the first stage was trained on the LUng Nodule Analysis (LUNA) ‘16 dataset, which itself is a subset of the Lung Image Database Consortium (LIDC-IDRI) dataset [1]. The LUNA16 dataset includes 888 CT scans with slice thickness less than 2.5 mm, along with precise location information in world coordinates and diameter for a total of 1186 nodules *≥* 3 mm in diameter. For each of these nodules, we obtained corresponding malignancy scores assigned by four radiologists on scale of 1 to 5 (1 being benign and 5 malignant) from the LIDC-IDRI dataset. As negative examples, we used the extended candidate set provided by the LUNA16 dataset for the ‘false positive reduction’ track, and assigned them malignancy scores of 0.

#### Training and Evaluation

##### Image preprocessing

Given a scan, the pixel intensities are converted to standard Hounsfield units (HU). Next, a window level of *−*400 HU and window width of 1600 HU is applied. Finally, the CT volume is resampled to isotropic 1 mm *×* 1 mm *×* 1 mm voxel dimensions.

##### Training the nodule detector

The CNN model for nodule detection and malignancy prediction was trained on the LUNA16 dataset. After CT image preprocessing, we extracted 3D cube patches to be input to the CNN. During training, the number of negative samples far exceeded the number of positive samples (*≈* 400 : 1); therefore, we used majority downsampling to create balanced training set. We used binary crossentropy and mean absolute error losses for the nodule probability and malignancy outputs respectively. We used random translations and random flips for data augmentation during training. The model was trained using stochastic gradient descent with step decay for the learning rate. We used a 80 : 20 train-validation split, and monitored the validation loss, saving the best models. The training was run for 20 epochs without any early-stopping.

##### Nodule detection for the NLST cohort

The trained CNN model is then used to generate nodule candidates for the NLST cohort. After preprocessing the CT scans, we used a 3D sliding window with a step size of 12 pixels in x, y and z directions, extracting 3D patches and feeding it to the CNN. If the predicted nodule probability exceeded a chosen threshold (0.6 in our case), we recorded the the predicted nodule probability and malignancy and treated the center of the 3D patch as the predicted nodule location.

##### Cancer prediction

For training the xgboost model for cancer prediction, as described we used the predicted nodule information as input and binary cancer vs no-cancer labels as output. For evaluation, we used a 50 : 50 train-test split and evaluated performance on the testing split. We repeated this for 1000 random splits to obtain a robust estimate of the performance metrics we report here.

## 3 Results

Our cancer prediction model shows significant performance as summarized in Table 1. Each row summarizes the performance metrics of the predictor obtained at the corresponding time-point. Note that the predictor at a given time-point uses the scans at that and all previous time-points for prediction. Fig. 1 shows the ROC and the precision-recall curves for our predictor at T2 (which uses the scans at all three time-points T0, T1 and T2). A key observation is the improvement in performance as more time-points are included for the prediction. For example, as shown in Fig. 2(b), the AUC increases from 0.75 at T0, to 0.80 at T1 to 0.85 at T2. Similar observations hold for the other performance metrics as well. Fig. 2(a) shows the sensitivity of our predictors broken down by cancer years; for a given year, we consider the patients who were confirmed cancers by that year and evaluate the recall of the predictor among these patients. As expected, for each predictor, the sensitivity decreases as it is, in general, more difficult to predict cancers that is diagnoses at the farther point in time. A second observation is the fact the sensitivity of the predictor at T1 for year *t* is greater than the sensitivity of the predictor at T0 for year *t* − 1, for *t* = −1, …, 6. The same holds for the predictors at time-points T2 and T1. This suggests that there is benefit to combining scans from more than one time-point for prediction, even if the scan time-points are in the past.

**Table 1:** Performance of our model (PPV: Positive predictive value, NPV: Negative predictive value). The 95% confidence intervals of the form (mean-delta, mean+delta) are shown as mean (delta) in the table.

| Time-point | AUC | Sensitivity | Specificity | PPV | NPV |
| --- | --- | --- | --- | --- | --- |
| T0 | 0.75 (0.03) | 0.61 (0.05) | 0.78 (0.06) | 0.72 (0.05) | 0.70 (0.03) |
| T1 | 0.80 (0.03) | 0.67 (0.06) | 0.80 (0.06) | 0.76 (0.05) | 0.72 (0.03) |
| T2 | 0.85 (0.02) | 0.75 (0.05) | 0.84 (0.04) | 0.82 (0.03) | 0.78 (0.03) |

**Fig. 1:**
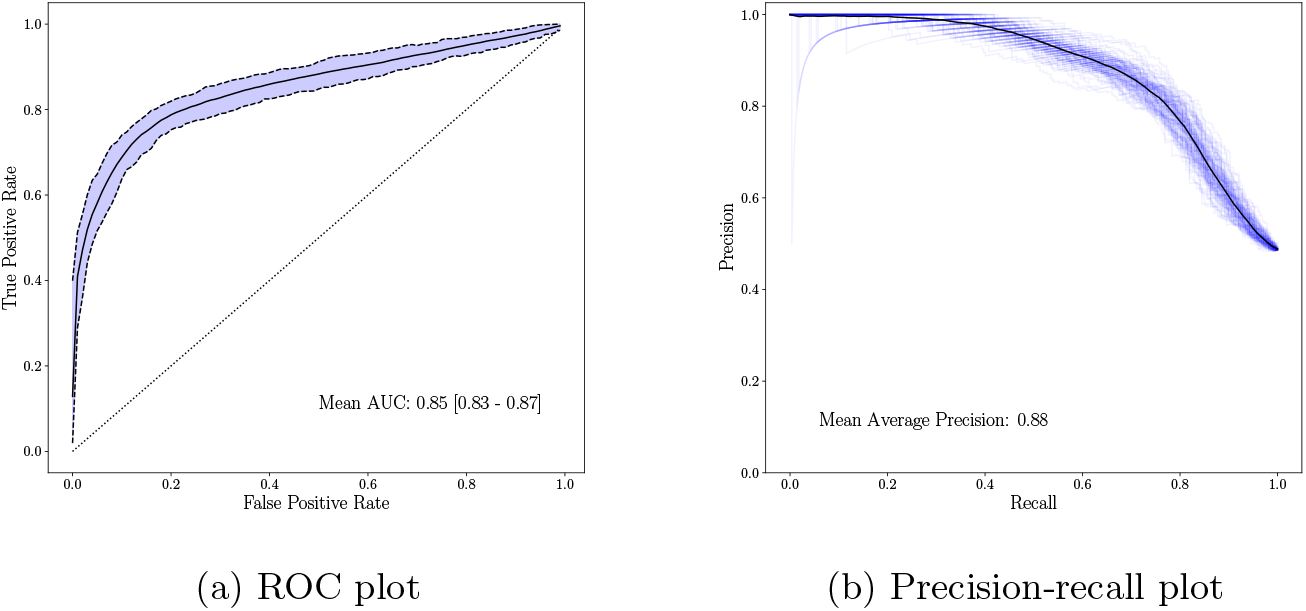
ROC and PR plots for predictor based on all three time-points, obtained using 1000 independent 50: 50 train-validation splits.

**Fig. 2:**
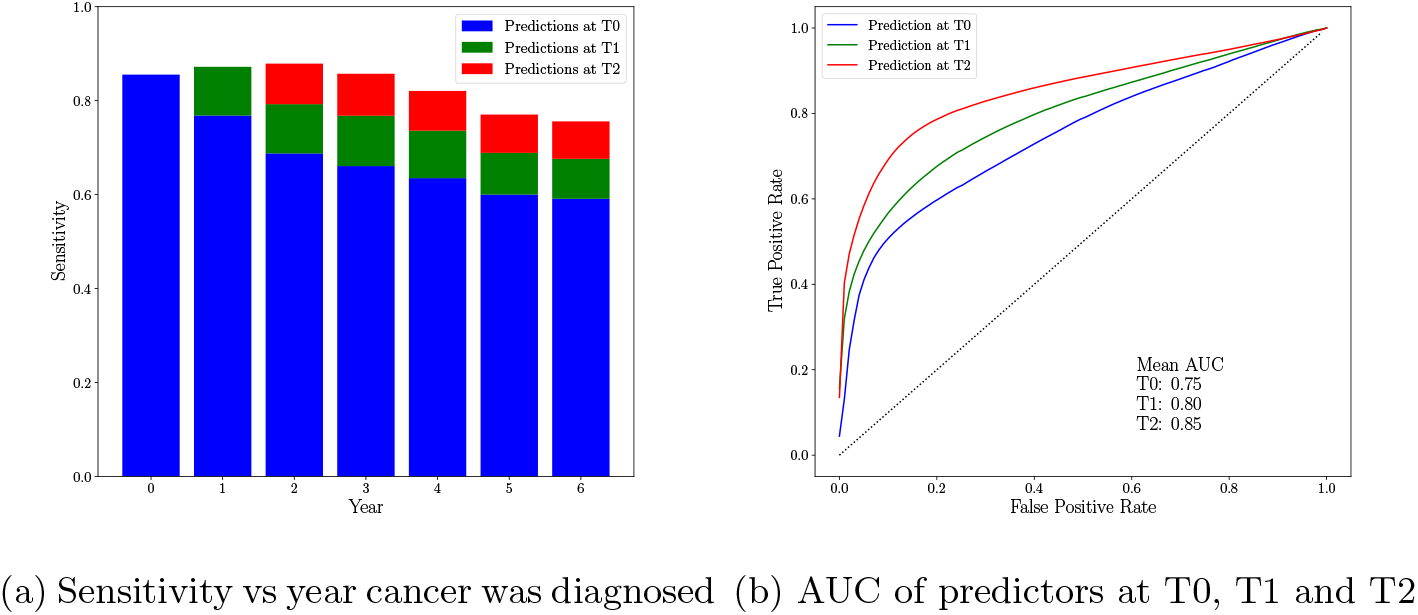
Performance comparison of predictors at T0, T1 and T2

We also built clinical only models based on the following features: age, gender, smoke-years, pack-years, race and ethnicity. We experimented with both tree based models such as xgboost and random forests, as well as linear models like logistic regression, and used a 10 fold crossvalidation strategy to evaluate performance. None of the models were successful in predicting cancer, with average cross-validated test AUCs around 0.5 in each case. This is not unexpected, as our cohort was chosen while controlling for these features.

## 4. Discussion

In this study, we have shown that it is possible to predict whether a patient with lung nodules *>* 4 mm has or will develop cancer in subsequent years based on lung CT scans only. The first stage of our model was trained on LIDC-IDRI dataset to detect nodules and predict their malignancy scores in the NLST dataset. Next, these nodule characteristics were combined in a xgboost model to predict cancer on a per patient basis. This showed that using up to three scans had the best performance with AUC of 0.85 to predict cancer.

Our study has limitations. We used only the lung CTs for predicting cancer; indeed, we controlled for other demographic and clinical features such as age, sex, and smoking history, which themselves may be predictive of cancer. Doing so allowed us to show that the lung CTs alone are predictive of cancer, lending credence to our hypothesis that features in the nodule or lung tissue may predict cancer, even years before it is confirmed. However, in a clinical setting, we would likely want to incorporate predictive demographic and clinical features as well, and that may improve the predictive performance as well. Additionally, in the second stage of our model, when multiple CT sequences are present in a single time-point, we chose one of them at random, and used its nodule information predicted by the first stage, for prediction of cancer. We did this without taking into account the variability of CT acquisition parameters such as convolution kernel, and we did not assess the impact of this variability on the prediction performance. This heterogeneity across CT scans may have degraded performance of nodule detection in the first stage of our model as well.

Finally, our current model just predicts cancer on a per patient basis, without pinpointing which nodules were most responsible for the prediction. In the future, we plan to incorporate such per nodule analysis in our model to improve its interpretability and trustworthiness for clinical usage.

## Data Availability

We used publicly available data which can be found here:

NLST: https://wiki.cancerimagingarchive.net/display/NLST/National+Lung+Screening+Trial

LIDC: https://wiki.cancerimagingarchive.net/pages/viewpage.action?pageId=1966254

https://wiki.cancerimagingarchive.net/display/NLST/National+Lung+Screening+Trial

https://wiki.cancerimagingarchive.net/pages/viewpage.action?pageId=1966254

